# Age-adjusted Charlson comorbidity index score is the best predictor for severe clinical outcome in the hospitalized patients with COVID-19 infection: a result from nationwide database of 5,621 Korean patients

**DOI:** 10.1101/2020.10.26.20220244

**Authors:** Do Hyoung Kim, Hayne Cho Park, Ajin Cho, Juhee Kim, Kyu-sang Yun, Jinseog Kim, Young-Ki Lee

**Affiliations:** Department of Internal Medicine, Kangnam Sacred Heart Hospital, Hallym University College of Medicine, Seoul, Korea; Hallym University Kidney Research Institute, Seoul, Korea; Department of Bigdata and Applied Statistics, Dongguk University, Gyeongju, Korea

## Abstract

Aged population with comorbidities demonstrated high mortality rate and severe clinical outcome in the patients with coronavirus disease 2019 (COVID-19). However, whether age-adjusted Charlson comorbidity index score (CCIS) predict fatal outcomes remains uncertain. This retrospective, nationwide cohort study was performed to evaluate patient mortality and clinical outcome according to CCIS among the hospitalized patients with COVID-19 infection. We included 5,621 patients who had been discharged from isolation or had died from COVID-19 by April 30, 2020. The primary outcome was composites of death, admission to intensive care unit (ICU), use of mechanical ventilator or extracorporeal membrane oxygenation. The secondary outcome was mortality. Multivariate Cox proportional hazard model was used to evaluate CCIS as the independent risk factor for death. Among 5,621 patients, the high CCIS (≥3) group showed higher proportion of elderly population and lower plasma hemoglobin and lower lymphocyte and platelet counts. The high CCIS group was an independent risk factor for composite outcome (HR 3.63, 95% CI 2.45-5.37, *P* < 0.001) and patient mortality (HR 22.96, 95% CI 7.20-73.24, *P* < 0.001). The nomogram demonstrated that the CCIS was the most potent predictive factor for patient mortality. The predictive nomogram using CCIS for the hospitalized patients with COVID-19 may help clinicians to triage the high-risk population and to concentrate limited resources to manage them.

## INTRODUCTION

The coronavirus disease 2019 (COVID-19) has been declared as a pandemic by the World Health Organization in March 2020^1^, and as of July 31, 2020, 16,864,828 people have been affected by the virus and 663,580 deaths were reported worldwide^2^. The Korean Centers for Disease Control and Prevention (KCDC) and relevant organization prepared policies for management for the COVID-19 outbreak as soon as the first case of COVID-19 was reported in Korea^3^. The Korean government not only performed extensive confirmatory tests for those who are suspected to have the disease but also implemented strict social distancing to prevent further transmission of disease. As the results, mild cases among young and healthy population were diagnosed as COVID-19 by screening tests but subsequent case fatality rate revealed lower than those reported worldwide. As of July 31, there were 14,269 confirmed cases of COVID-19 and 300 deaths in Korea (case fatality rate of 2.1% in Korea vs. 3.9% in average worldwide)^4^.

Nevertheless, there were some population with higher morbidity and mortality during COVID-19 outbreak. The situation reports from KCDC demonstrated that higher mortality among the patients with advanced age^4^. It is reasonable to think that the patients with advanced age show higher mortality because of reduced immunity and higher comorbidities such as diabetes or chronic kidney disease. Previous reports suggested old age and multiple comorbidities are independently associated with higher mortality^5-7^. However, most of the studies demonstrated individual category of disease such as diabetes or coronary heart disease as a risk factor but not weighted the effect of age and severity of multimorbidity as predictors of poor clinical outcomes.

Age-adjusted Charlson comorbidity index score (CCIS) has been developed and validated to predict the risk of mortality for use in longitudinal studies^8^. The score can be calculated from a weighted index consisted of age and the number and seriousness of comorbid diseases. The CCIS has been widely used to predict 10-year mortality among hospitalized patients^8^. The CCIS also has been validated in a various disease conditions such as acute stroke^9,10^, leukemia^11^, end stage renal disease^12,13^, and hip fracture^14,15^.

Recently, KCDC released COVID-19 database to the public. The database contains the clinical and epidemiologic data of 5,628 confirmed cases as well as their clinical outcome until April 30, 2020. We evaluated patient mortality and clinical outcome according to CCIS among the hospitalized patients with COVID-19 in Korea.

## RESULTS

### Baseline characteristics of the Korean COVID-19 cohort

The baseline characteristics of 5,621 patients are shown in Table 1. The mean duration from diagnosis to death or release from isolation was 25.6 days. The patients in 50s was the largest age group followed by those in 20s (Figure 1). The 3,304 (58.8%) patients were female. Female was predominant throughout all age groups except those younger than 20. Hypertension (21.3%) and diabetes mellitus (12.3%) were the most common two comorbidities. It was followed by dementia (4.2%), chronic heart disease (3.2%), malignancy (2.5%), asthma (2.3%), chronic liver disease (1.5%), congestive heart failure (1.0%), chronic kidney disease (1.0%), chronic obstructive pulmonary disease (0.7%), and connective tissue disease (0.7%) (Figure 1). The patients with low systolic blood pressure (BP) (<120mmHg) and diastolic BP (<80mmHg) were 1317 (24.0%) and 2113 (38.5%), respectively. Among 5621 confirmed cases, asymptomatic infection comprised of 25.8%. Among symptomatic patients, three most common symptoms were cough (41.7%), sputum (28.8%), and febrile sense (23.2%).

**Figure 1.**
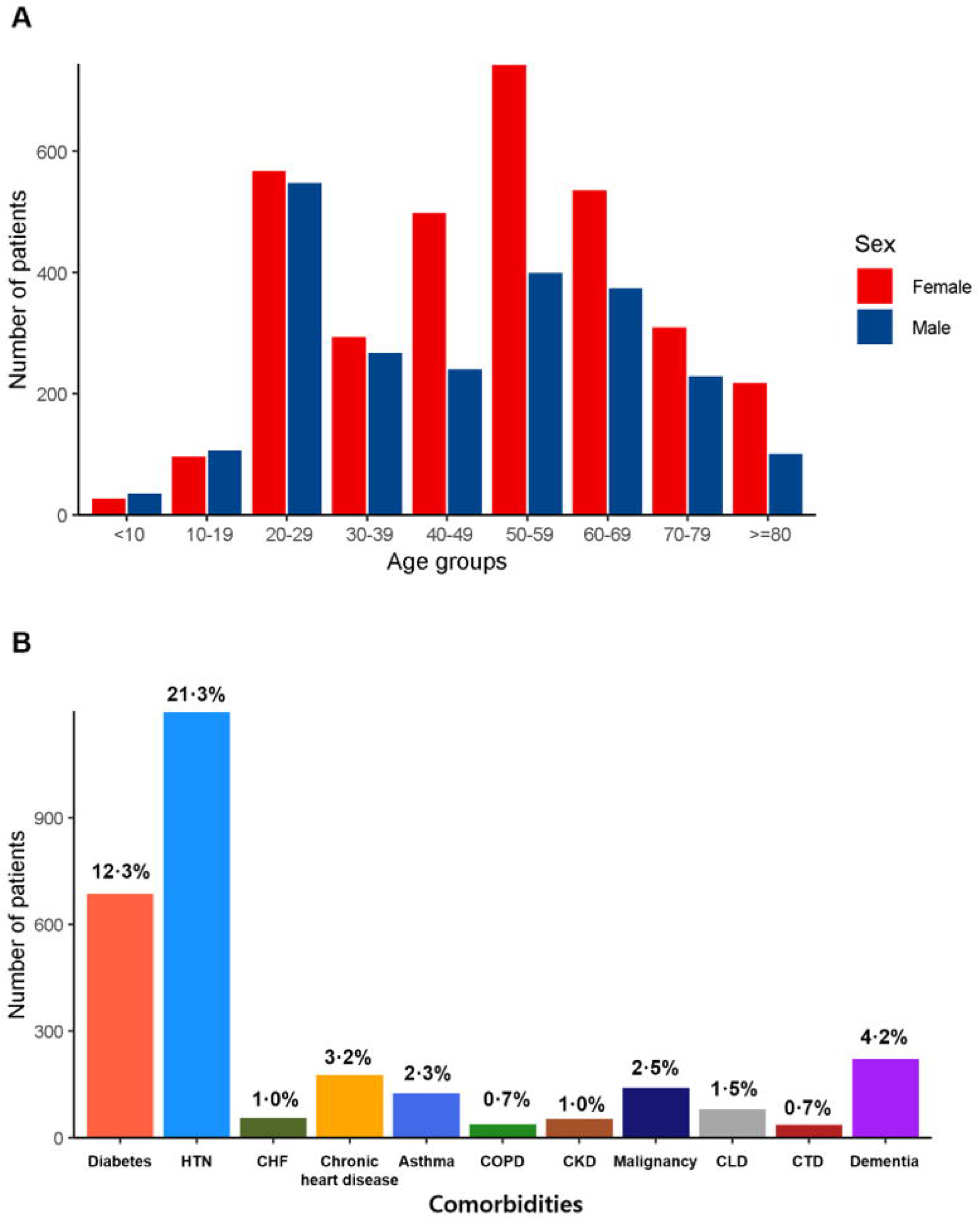
Demographic distribution of the patients with COVID-19 in Korea. A. Age and sex distribution; B. Proportion of underlying comorbidities. The patients in 50s was the largest age group followed by those in 20s. Female was predominant throughout all age groups except those younger than 20. Hypertension (21.3%) and diabetes mellitus (12.3%) were the most common two comorbidities. It was followed by dementia (4.2%), chronic heart disease (3.2%), malignancy (2.5%), asthma (2.3%), chronic liver disease (1.5%), congestive heart failure (1.0%), chronic kidney disease (1.0%), chronic obstructive pulmonary disease (0.7%), and connective tissue disease (0.7%) *Abbreviations*. CHF, Congestive heart failure; CKD, Chronic kidney disease; CLD, Chronic liver disease; COPD, Chronic obstructive pulmonary disease; COVID-19, coronavirus disease 2019; CTD, Connective tissue disease; HTN, hypertension.

The median score of CCIS was 2 (IQR, 0-3). The number of patients in CCIS score 0, 1-2, 3-4, ≥5 group was 1,890 (33.6%), 1,791 (31.9%), 1,399 (24.9%), and 541 (9.6%), respectively. Compared to the low CCIS group (<3), the high CCIS group (≥3) showed higher proportion of elderly patients and medical comorbidities. Presence of dyspnea, altered mentality, and nausea/vomiting at admission were more frequent in the high CCIS group. In addition, the number of lymphocyte and platelet count and plasma hemoglobin (Hb) level were significantly lower in high CCIS group compared to low CCIS group.

### Clinical outcomes according to CCIS

Composite events occurred in 368 patients (6.6 %) during the follow up period including 234 (4.2 %) deaths, 187 (3.3 %) admission cases of intensive care unit (ICU), 52 (0.9 %) cases of mechanical ventilation, and 11 (0.2%) cases of extracorporeal membrane oxygenation (ECMO) application. The composite outcomes occurred more frequently in high CCIS groups showing 178 patients (32.9 %) in CCIS ≥5 followed by 135 (9.7%) in CCIS 3-4, 36 (2.0%) in CCIS 1-2, and 19 (1.0%) in CCIS 0 (Table 2). The mortality rate was also higher in high CCIS groups demonstrating 154 patients (28.5%) in CCIS ≥5 followed by 68 (4.9%) in CCIS 3-4, 11 (0.6%) in CCIS 1-2, and 1 (0.1%) in CCIS 0 (Figure 2).

**Figure 2.**
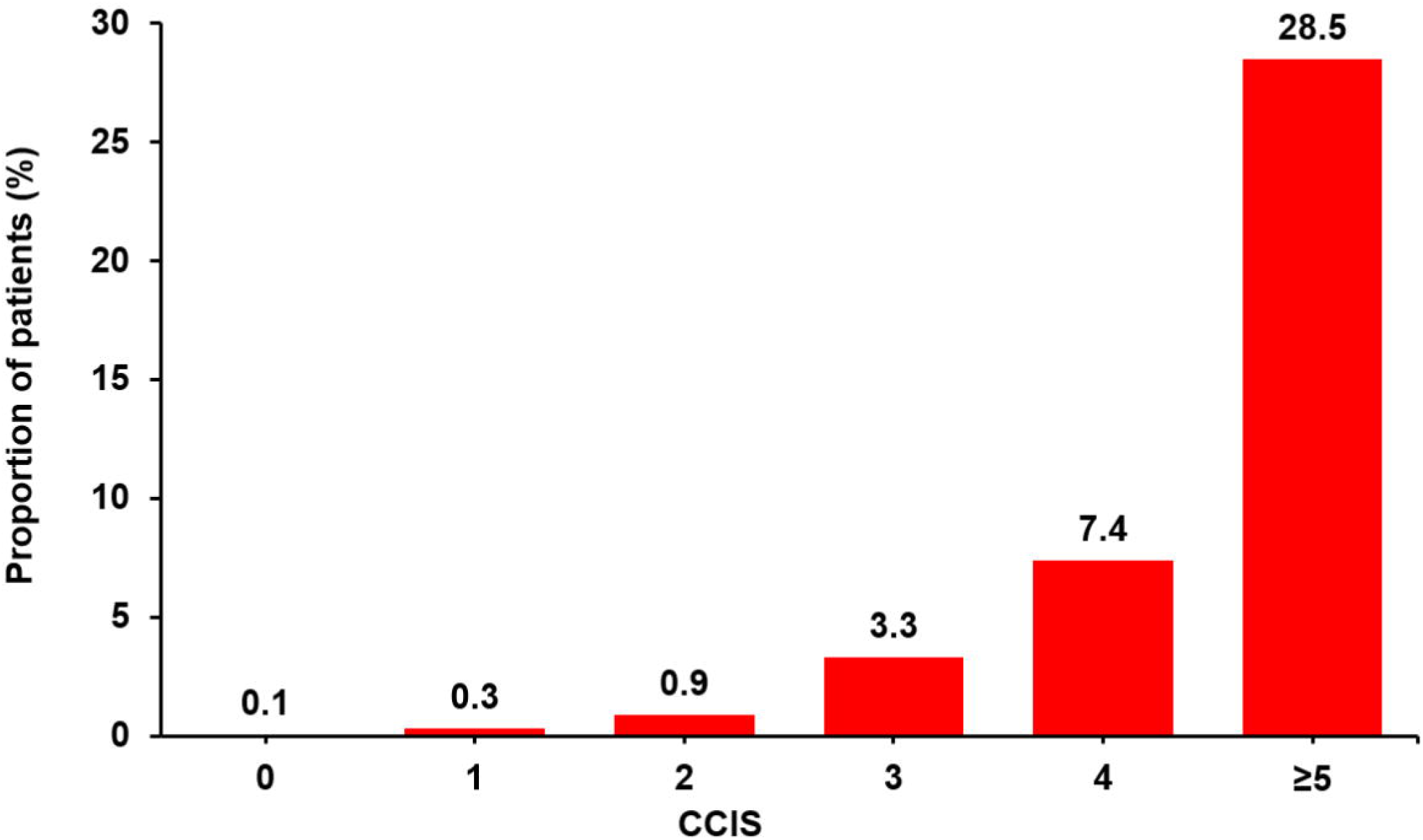
Proportion of death according to CCIS of patients with COVID-19. The morality rate increased as CCIS increases. The mortality rate was 28.5% in CCIS ≥5 followed by 4.9% in CCIS 3-4, 0.6% in CCIS 1-2, and 0.1% in CCIS 0. *Abbreviations*. CCIS, age-adjusted Charlson comorbidity index score; COVID-19, coronavirus disease 2019.

### Risk factors for severe clinical outcomes

We performed multivariate Cox proportional analysis to find out high CCIS ≥3 was an independent risk factor for composite outcome (hazard ratio (HR), 3.63 [95% confidence interval (CI), 2.45-5.37], P < 0.001) together with male sex (HR, 1.76 [95% CI 1.32–2.34], P < 0.001), body mass index (BMI) <18.5 kg/m^2^ (HR, 2.36 [95% CI 1.49–3.75], P < 0.001), presence of dyspnea (HR, 2.88 [95% CI 2.16–3.83], P < 0.001), lymphopenia (HR, 2.15 [95% CI 1.59-2.91], P < 0.001), and anemia (HR, 1.80 [95% CI 1.33-2.43], P < 0.001) (See Figure 3 and Supplementary Table S1 online).

**Figure 3.**
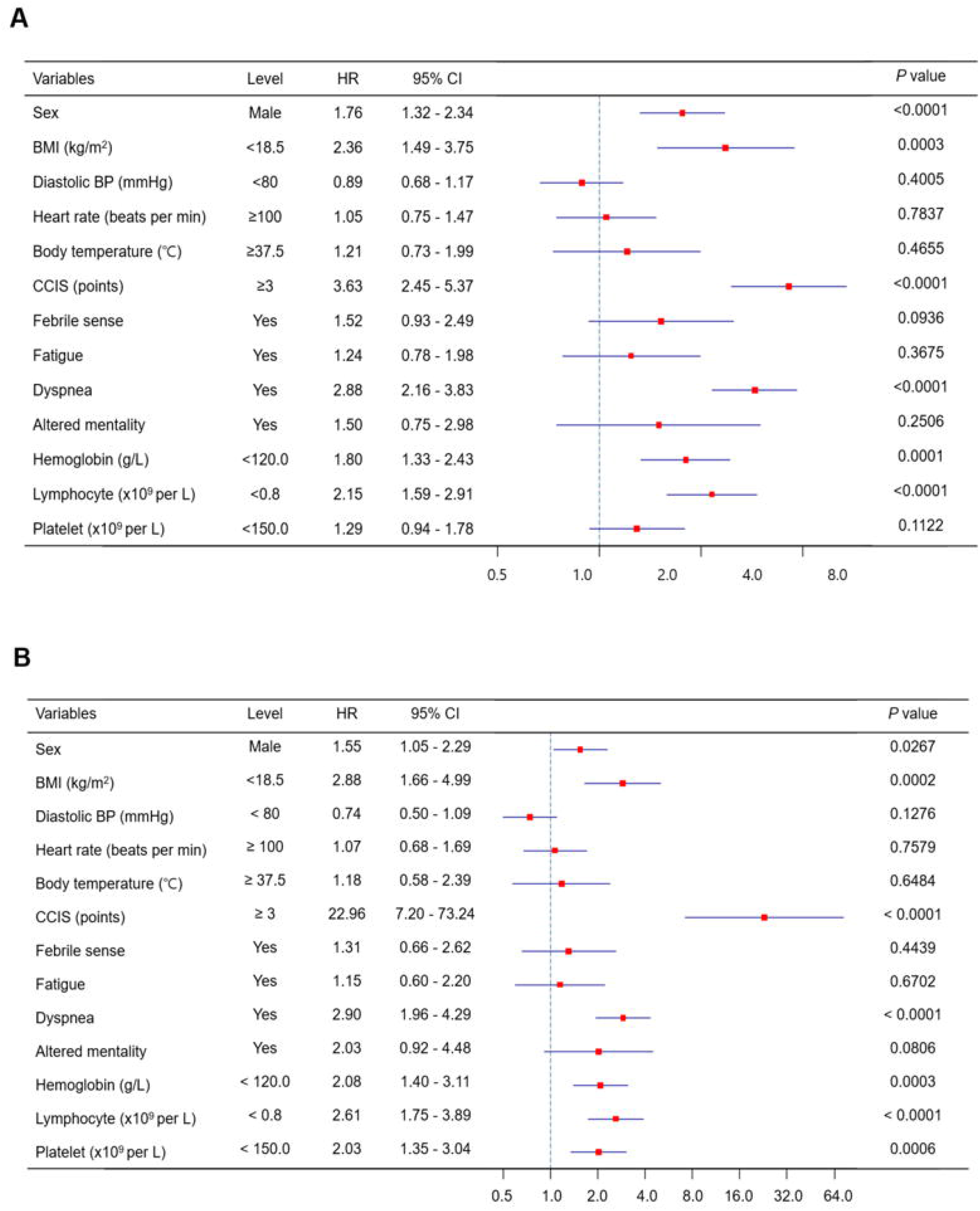
Risk factor for severe clinical outcomes in the patients with COVID-19. A. Risk factors for the composite outcome; B. Risk factors for patient mortality. Multivariate Cox proportional hazard model demonstrated CCIS is an independent risk factor for composite outcome and mortality after adjusting other risk factors. *Abbreviations*. BMI, body mass index; BP blood pressure; CCIS, age-adjusted Charlson comorbidity index score; CI, confidence interval; COVID-19, coronavirus disease 2019.

Then, we analyzed the risk factors for mortality (see Supplementary Figure S1 online). The patients older than 70 years old demonstrated higher mortality risk compared younger patients. The patients with high CCIS ≥3 also showed higher risk for mortality compared to low CCIS group. When we assessed the risk factors for mortality in univariate analysis, old age ≥70 years, male sex, high CCIS ≥3, anemia, lymphopenia, and thrombocytopenia were significantly associated with higher mortality rate. In multivariate Cox proportional analysis, risk of mortality was higher in patients with CCIS ≥3 (HR, 22.96 [95% CI 7.20 - 73.24], P < 0.001), male sex (HR, 1.55 [95% CI 1.05–2.29], P = 0.027), BMI < 18.5 kg/m^2^ (HR, 2.88 [95% CI 1.66–4.99], P < 0.001), presence of dyspnea (HR, 2.90 [95% CI 1.96–4.29], P < 0.001), lymphopenia (HR, 2.61 [95% CI 1.75-3.89], P < 0.001), anemia (HR, 2.08 [95% CI 1.40-3.11], P < 0.003), and thrombocytopenia (HR, 2.03 [95% CI 1.35–3.04], P < 0.001) (Figure 3).

### Prediction model for case fatality

The predictive nomogram was constructed based on the multivariate Cox analysis for mortality. To calculate 14-, and 28-day overall survival probability was calculated from the summed points of sex, BMI, CCIS, presence of dyspnea, plasma Hb, lymphopenia, and thrombocytopenia (Figure 4). The nomogram showed that CCIS was the most important factor contributing to the prognosis followed by the presence of dyspnea, low BMI <18.5 kg/m2, lymphopenia (<0.8 x10^9^/L), thrombocytopenia (<150.0 x10^9^/L), anemia (<12.0 g/dL), and male sex. The Harrell concordance index (C-index) value for prediction of overall survival was 0.933, and R^2^ value was 0.991 in 14-day and 0.990 in 28-day prediction model (see Supplementary Figure S2 online).

**Figure 4.**
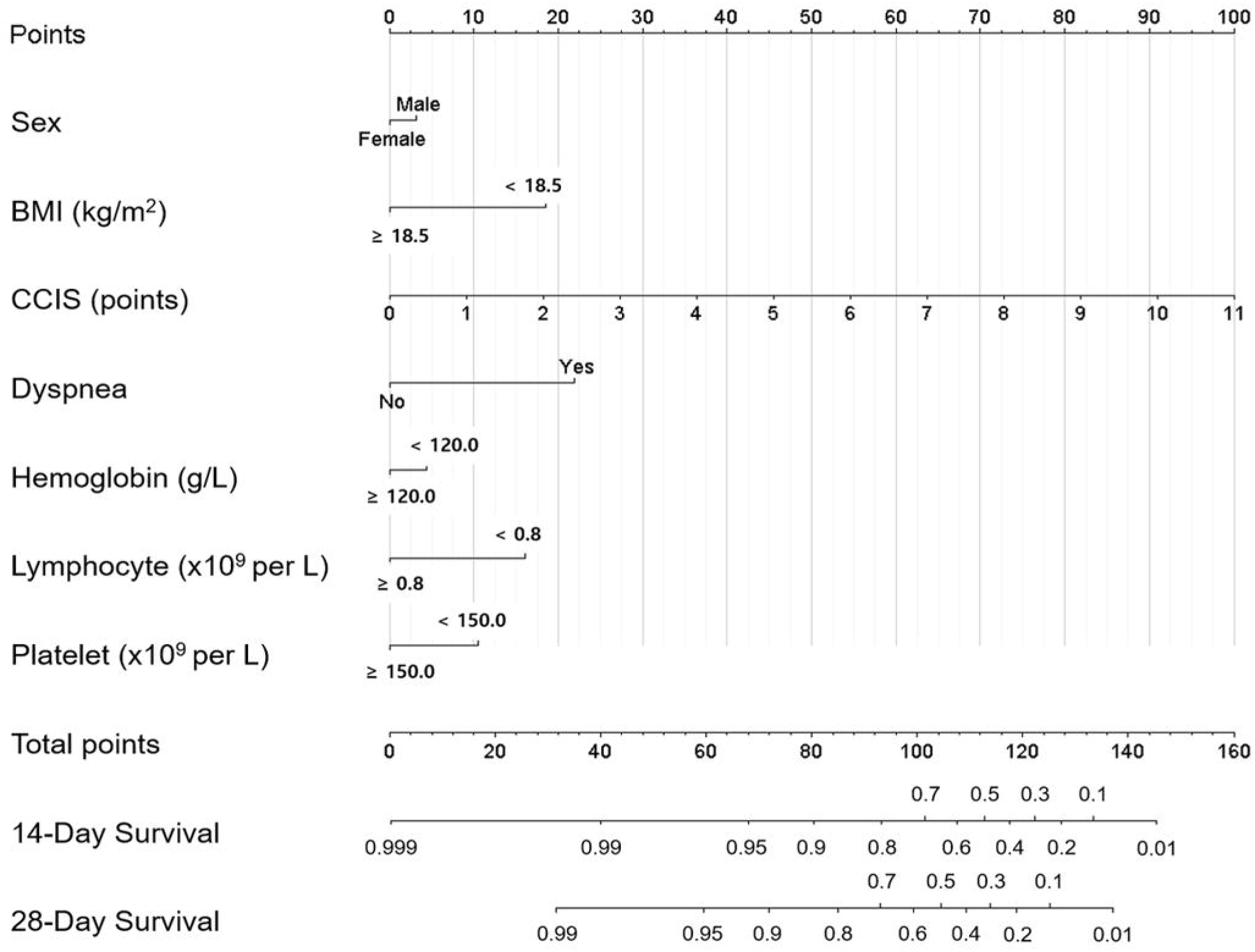
Prognostic nomogram for prediction of the overall survival probability of patients with COVID-19. The nomogram demonstrates CCIS is the potent predictor for 14-day and 24-day survival of the patients. *Abbreviations*. BMI, body mass index; CCIS, age-adjusted Charlson comorbidity index score; COVID-19, coronavirus disease 2019.

## DISCUSSION

In this study, we described the effect of age and comorbidity upon composites of severe clinical outcomes (death, ICU admission, mechanical ventilation, or ECMO) among the hospitalized patients with COVID-19. The patients with high CCIS showed older age, lower plasma level of Hb, and lower lymphocyte and platelet counts. The high CCIS (≥3) group was an independent risk factors for composite outcome (HR 3.63, 95% CI 2.45-5.37, p<0.0001) and patient mortality (HR 7.05, 95% CI 2.13-23.33, p=0.0014).

Korean COVID-19 cohort demonstrated large proportion of younger population with age <50 (34.7%) and asymptomatic confirmed cases (25.7%). The patients without any comorbidities (CCIS 0) comprised of 33.1% of total population. Median CCIS was 2 for the Korean COVID-19 cohort. Previous report from Wuhan, China demonstrated higher proportion of younger population with age <50 (56.0%) as well as greater proportion of the patients without any comorbidities (76.3%)^16^. The brief report from Danish population also demonstrated that higher proportion of confirmed cases without any comorbidities (CCIS 0, 65.0%)^17^. On the other hands, previous papers from United States of America demonstrated higher proportion of older people (mean age of 61.0) and lower proportion of the patients without existing comorbidities (27.4%)^18^. In Italy where one of the highest case fatality rates were reported, the non-survivor demonstrated advanced age (79.6±0.9 years vs. 64.7±0.4 years, p=0.0001) and higher CCIS (4.3±0.15 vs. 2.6±0.05, p=0.0001) compared to survivors^5^. The observed differences of baseline characteristics among various countries may be related not only to patient demographics but also the degree of performing active screening tests against asymptomatic population.

Our study is the first large nationwide cohort study to demonstrate that CCIS is the best predictor for severe clinical outcome in the patients with COVID-19. Previous paper by Chen et al. demonstrated that advanced age is the strongest risk factor for a fatal outcome^6^. In addition to advanced age, preexisting comorbid conditions including coronary heart disease and cerebrovascular disease, shortness of breath, and levels of procalcitonin and aspartate aminotransferase were related to fatal outcome. However, previous study by Zhou et al. presented only advanced age but not comorbidity was an independent risk factor for mortality^19^. Another study by Ji et al. also evaluated the risk factors for progression risk of pneumonia in COVID-19 patients^20^. They showed that presence of comorbidities and advanced age both contribute to higher risk of disease progression. Korean group also recently published the paper about the impact of comorbidities upon the mortality^7,21^. However, these studies did not use CCIS to calculate the weight of the comorbidities. There were several studies suggesting CCIS as an independent risk factor for hospitalization or patient mortality among the patients with COVID-19^17,18,22^. However, to our knowledge, there was no study showing prediction model of fatal outcome using CCIS.

In the same vein with previous studies, the advanced age >50 years old demonstrated high risk of mortality and ICU admission. The previous study performed by Sung et al. showed relatively young patients under age 50 recovered without oxygen therapy or ICU care^23^. On the other hands, the patients of ≥50 years of age demonstrated high case fatality rate reaching 14% in the patients over 80 years old. Our study also confirmed that advanced age itself is an independent factor for composites of severe clinical outcome.

There are some limitations to our study. First, the nationwide database from KCDC only offered limited data. For example, there are no information about chest radiograph findings or laboratory findings such as C-reactive protein or serum creatinine. Therefore, we could not adjust potential risk factors in our study. Second, due to the nature of survey without collecting actual previous medical history from ICD-10 diagnosis code, some information about comorbidities are missing. Therefore, we modified some categories of chronic illness included in the original CCIS. For example, we scored all forms of coronary artery disease including myocardial infarction and chronic heart disease with a score of 1. Previous report also demonstrated the usefulness of modified form of CCIS in prediction of clinical outcome in renal patients^24^. In addition, the information about some chronic conditions such as peripheral vascular disease, cerebrovascular disease or peptic ulcer disease were missing. We calculated CCIS based on the available data excluding those categories of missing data. Therefore, CCIS in our cohort may be underestimated than actual CCIS. Lastly, there can be ethnic or racial difference in clinical outcomes of COVID-19. Therefore, our result may not be applicated to the different ethnicity or population. However, recent paper suggested that racial difference did not contribute to different clinical outcome^22^. Nevertheless, there should be a validation test in each population using our predictive tool before clinical application.

In conclusion, our study provided convenient predictive tool using CCIS for the hospitalized patients with COVID-19 to calculate the risk of severe clinical outcome at admission. The CCIS together with initial symptom and laboratory findings may lead us to triage the patients most likely to progress and concentrate our efforts to save their lives.

## METHODS

### Study design and participants

This is a retrospective cohort study using the nationwide COVID-19 database provided by KCDC. A total of 5,628 patients were included in the database who had been confirmed to have COVID-19 and had received treatment from 100 hospitals until April 30, 2020. Among them, 7 patients who were confirmed for COVID-19 after death were excluded, and a total of 5,621 patients were included in the analysis. According to the definition provided by the KCDC^25^, the confirmed case was defined as a patient who had been confirmed to be infected with COVID-19 by real-time reverse transcriptase-polymerase chain reaction assay or virus isolation from nasal and/or pharyngeal swab specimens regardless of the clinical symptoms. The study protocol was reviewed and approved by the Institutional Review Board of the Kangnam Sacred Heart Hospital, Seoul, Korea (HKS 2020-06-025). The informed consent was waived due to retrospective nature of the study.

### Data collection

Clinical and epidemiological data were collected in a retrospective manner and were anonymized before release. Demographic data including age by decade, sex, survival status, and duration of isolation were collected. Information about whether the patient was admitted to ICU or applied mechanical ventilator or ECMO were also collected. The BMI, systolic and diastolic BP, heart rate, and body temperature at initial visit were also measured and reported. The clinical symptoms associated with COVID-19 were reported including febrile sense, cough, sputum, sore throat, rhinorrhea, myalgia, shortness of breath, headache, altered consciousness/confusion, nausea or vomiting, diarrhea. Laboratory assessments consisted of plasma Hb, white blood cell, lymphocyte, and platelet count. The categories of comorbidities were assessed including diabetes mellitus, hypertension, heart failure, chronic heart disease, asthma, chronic obstructive pulmonary disease, chronic kidney disease, malignancy, chronic liver disease, connective tissue disease, and dementia. The CCIS was calculated using the method described in the previous study^8,24^. Due to lack of available data, all kinds of chronic heart disease were considered to have congestive heart failure and all forms of chronic liver disease were considered to have mild liver disease. The presence of peripheral vascular disease, cerebrovascular disease, peptic ulcer disease, hemiplegia, end-stage renal disease, or acquired immunodeficiency syndrome could not be known from the current database. All kinds of malignancy were considered non-metastatic.

### Study outcomes

The primary endpoint was defined as the composite of death, admission to ICU, application of mechanical ventilation or ECMO. The secondary endpoints were patient mortality and ICU admission rate. The primary and secondary endpoints were compared between low and high CCIS groups. Finally, we established a prediction model for patient mortality through risk factor analysis.

### Statistical Analysis

The baseline characteristics and clinical outcomes were compared among CCIS groups. We divided the patients into 4 groups according to CCIS: CCIS 0, 1-2, 3-4, and ≥5. The normally distributed numerical variables were expressed as the mean ± standard deviation, whereas variables with skewed distributions were expressed as the median and interquartile range. Statistical comparisons between continuous variables were performed with an independent Student t-test or one-way analysis of variance for more than two group. For the data without normal distribution, the Wilcoxon Signed Rank Test for two groups or Kruskal Wallis Test for more than two groups were performed. The χ2 test and Fisher exact test were applied to categorical variables as appropriate.

The Kaplan-Meier method was used to compare composite events-free survival curves, and differences were assessed utilizing the log-rank test. We used univariate and multivariate Cox proportional hazard model to estimate risk factors associated with composite outcome and patient mortality. Age was excluded from the multivariate analysis because of its potential interaction with CCIS. We used univariate and multivariate logistic regression models to evaluate the risk factors for ICU admission or use of mechanical ventilation. We used categorical variables to assess independent risk factors for clinical outcomes as follows: age group of <50 (reference), 50-69, and ≥70 years, female (reference) vs. male, BMI ≥18.5 (reference) vs. <18.5 kg/m^2^, systolic BP ≥120 (reference) vs. <120 mmHg, diastolic BP ≥80 (reference) vs. <80 mmHg, heart rate <100 (reference) vs. ≥100 beats per minute, body temperature <37.5 (reference) vs. ≥37.5□, CCIS <3 (Reference) vs. ≥3 points, lymphocyte count≥0.8 (reference) vs. <0.8 x 10^9^/L, Hb≥12.0 (reference) vs. <12.0 g/dL, and platelet count ≥150.0 (reference) vs. <150.0 x 10^9^/L.

A nomogram to predict 14-day and 28-day mortality risk of the patient was built based on the variables found in multivariate Cox proportional hazard model. In the nomogram, CCIS was used as a variable as a continuous variable to check the impact per CCIS. The maximum score of each variable was set as 100. The performance of the nomogram was measured based on the C-index. The nomogram was validated in calibration plots with 1,000 bootstrap samples in which the estimated survival probability was compared with the observed value. All statistical analysis was performed by using R version 4.0.2 (R Foundation for Statistical Computing; http://www.r-project.org/). P value < 0.05 was considered statistically significant.

### Data Availability Statement

The data that support the findings of this study are available from KCDC, but restrictions apply to the availability of these data and so are not publicly available. Data are however available from the authors upon request and with permission of KCDC.

## Supporting information

Suppplementary Materials

## Acknowledgements

We acknowledge all the health-care workers involved in the diagnosis and treatment of COVID-19 patients in South Korea. We thank the Central Disease Control Headquarters, National Medical Center and the Health Information Manager in 100 hospitals for their effort in collecting the medical records. There was no specific funding for this work.

## Authors’contributions

All authors participated in the research and preparation of the manuscript. K.D.H. and P.H.C. equally contributed to this manuscript as 1^st^ authors who drafted this paper. P.H.C. and L.Y.K. contributed to the conception of the work and study design. K.J.S. contributed to statistical analyses and producing tables and figures. K.D.H., K.J., and Y.K.S. contributed to the acquisition of data and manipulating the data to the analyzable format. L.Y.K. had full access to all the data in the study and take responsibility for the integrity and accuracy of the data. C.A. and L.Y.K. supervised the work and revised the manuscript.

