## Supplementary material for "Age-adjusted Charlson comorbidity index score is the best predictor for severe clinical outcome in the hospitalized patients with COVID-19 infection: a result from nationwide database of 5,621 Korean patients": Suppplementary Materials

**Supplementary** **Table S1. Univariate and multivariate Cox proportional analysis of predictors associated with composite outcomes.**

|  | **Univariate** | | **Multivariate** | |
| --- | --- | --- | --- | --- |
|  | **Hazard ratio (95% CI)** | *P* **value** | **Hazard ratio (95% CI)** | *P* **value** |
| Age 50-69 years | 4.55 (3.01-6.89) | < 0.001 |  |  |
| ≥70 years | 22.09(14.91-32.74) | < 0.001 |  |  |
| Male | 1.70 (1.38-2.08) | < 0.001 | 1.76 (1.32-2.34) | < 0.001 |
| BMI <18.5 kg/m^2^ | 1.78 (1.15 – 2.76) | 0.01 | 2.36 (1.49-3.75) | < 0.001 |
| Systolic BP <120 mmHg | 1.05(0.82-1.35) | 0.673 |  |  |
| Diastolic BP <80 mmHg | 1.26(1.02-1.55) | 0.033 | 0.89 (0.68-1.17) | 0.401 |
| Heart rate ≥100 / min | 1.60 (1.26-2.03) | < 0.001 | 1.05 (0.75 - 1.47) | 0.784 |
| Body temperature ≥37.5 ℃ | 2.31(1.85-2.88) | < 0.001 | 1.21 (0.73 – 1.99) | 0.466 |
| CCIS ≥3 | 10.43(7.65-14.22) | < 0.001 | 3.63 (2.45-5.37) | < 0.001 |
| Diabetes | 3.03 (2.43-3.77) | < 0.001 |  |  |
| Hypertension | 3.92(3.19-4.82) | < 0.001 |  |  |
| Congestive heart failure | 5.58 (3.55-8.77) | < 0.001 |  |  |
| Chronic heart disease | 2.90(2.06-4.10) | < 0.001 |  |  |
| Asthma | 1.68(1.00-2.81) | 0.051 |  |  |
| COPD | 3.31(1.80-6.06) | < 0.001 |  |  |
| Chronic kidney disease | 4.66 (2.91-7.46) | < 0.001 |  |  |
| Malignancy | 2.21 (1.43-3.40) | < 0.001 |  |  |
| Chronic liver disease | 1.49 (0.77-2.89) | 0.240 |  |  |
| Connective tissue disease | 1.18 (0.38-3.66) | 0.781 |  |  |
| Dementia | 6.62 (5.14-8.52) | < 0.001 |  |  |
| Any symptom | 1.56 (1.18-2.06) | 0.002 |  |  |
| Febrile sense | 2.24 (1.82-2.75) | < 0.001 | 1.52 (0.93-2.49) | 0.094 |
| Fatigue | 1.89 (1.29-2.76) | 0.001 | 1.24 (0.78-1.98) | 0.368 |
| Dyspnea | 5.74 (4.67-7.06) | < 0.001 | 2.88 (2.16-3.83) | < 0.001 |
| Altered mentality | 13.85 (9.13-21.01) | < 0.001 | 1.50 (0.75-2.98) | 0.251 |
| Anemia (Hb <12.0 g/dL) | 2.71 (2.18-3.35) | < 0.001 | 1.80 (1.33-2.43) | < 0.001 |
| Lymphopenia  (lymphocyte counts <0.8 ×10⁹/L) | 6.44 (5.19-7.99) | < 0.001 | 2.15 (1.59-2.91) | < 0.001 |
| Thrombocytopenia  (platelet counts <150 ×10⁹/L) | 2.96 (2.36-3.73) | < 0.001 | 1.29 (0.94-1.78) | 0.112 |

BMI, body mass index; BP, blood pressure; CCIS, age-adjusted Charlson comorbidity index score; CI, confidence interval COPD, chronic obstructive pulmonary disease; Hb, hemoglobin.

**Supplementary Figure S1. Kaplan-Meier survival plots for different prognostic factors.** The figure displays the Kaplan-Meier survival plots according to (A) age, (B) CCIS, (C) sex, (D) lymphopenia (lymphocyte counts <0.8 ×10⁹/L), (E) anemia (Hb <12.0 g/dL), (F) thrombocytopenia (platelet counts <150.0 ×10⁹/L), (G) diabetes, (H) hypertension, (I) chronic heart disease, (J) chronic kidney disease, (K) malignancy, and (L) dementia. CCIS, age-adjusted Charlson comorbidity index score.

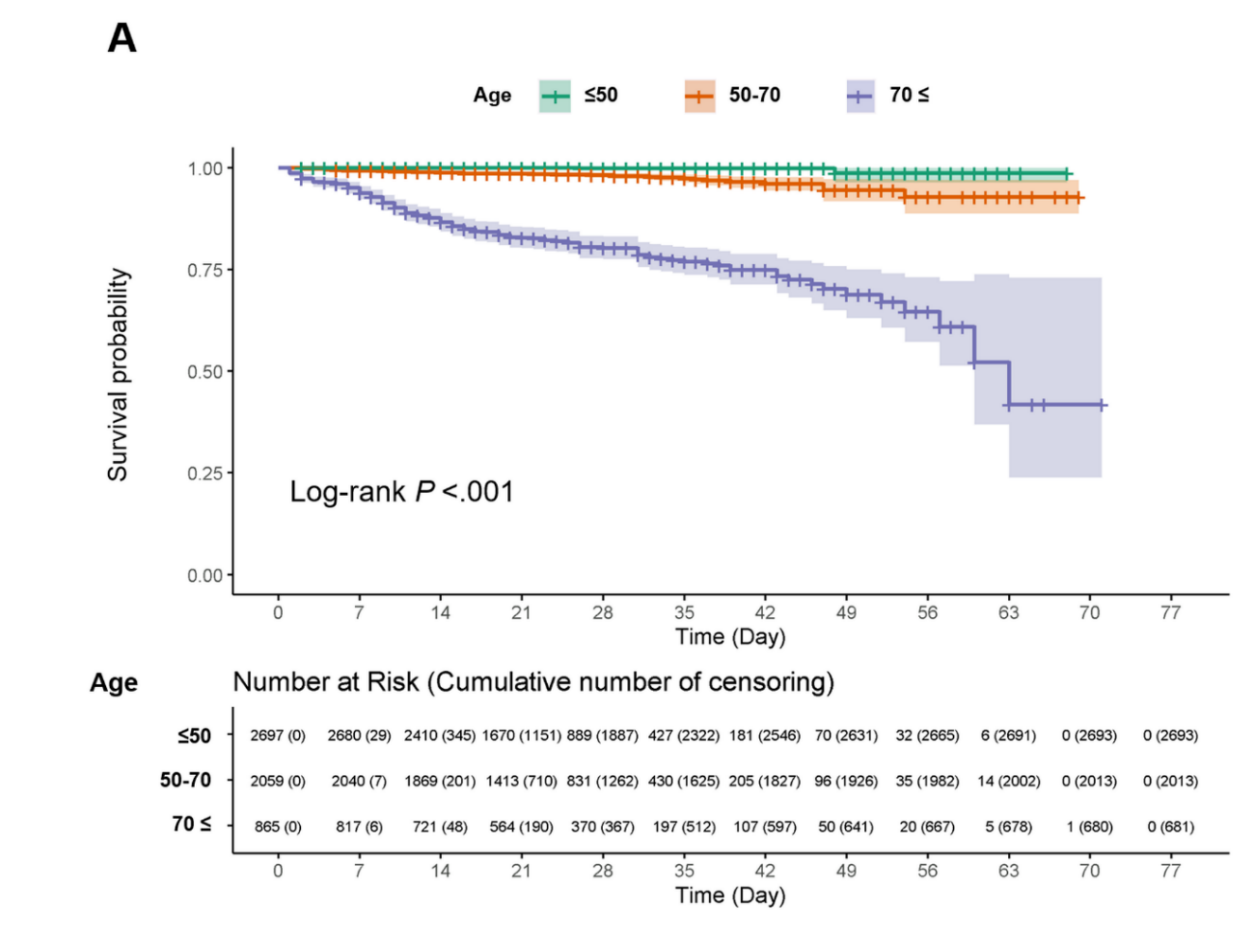

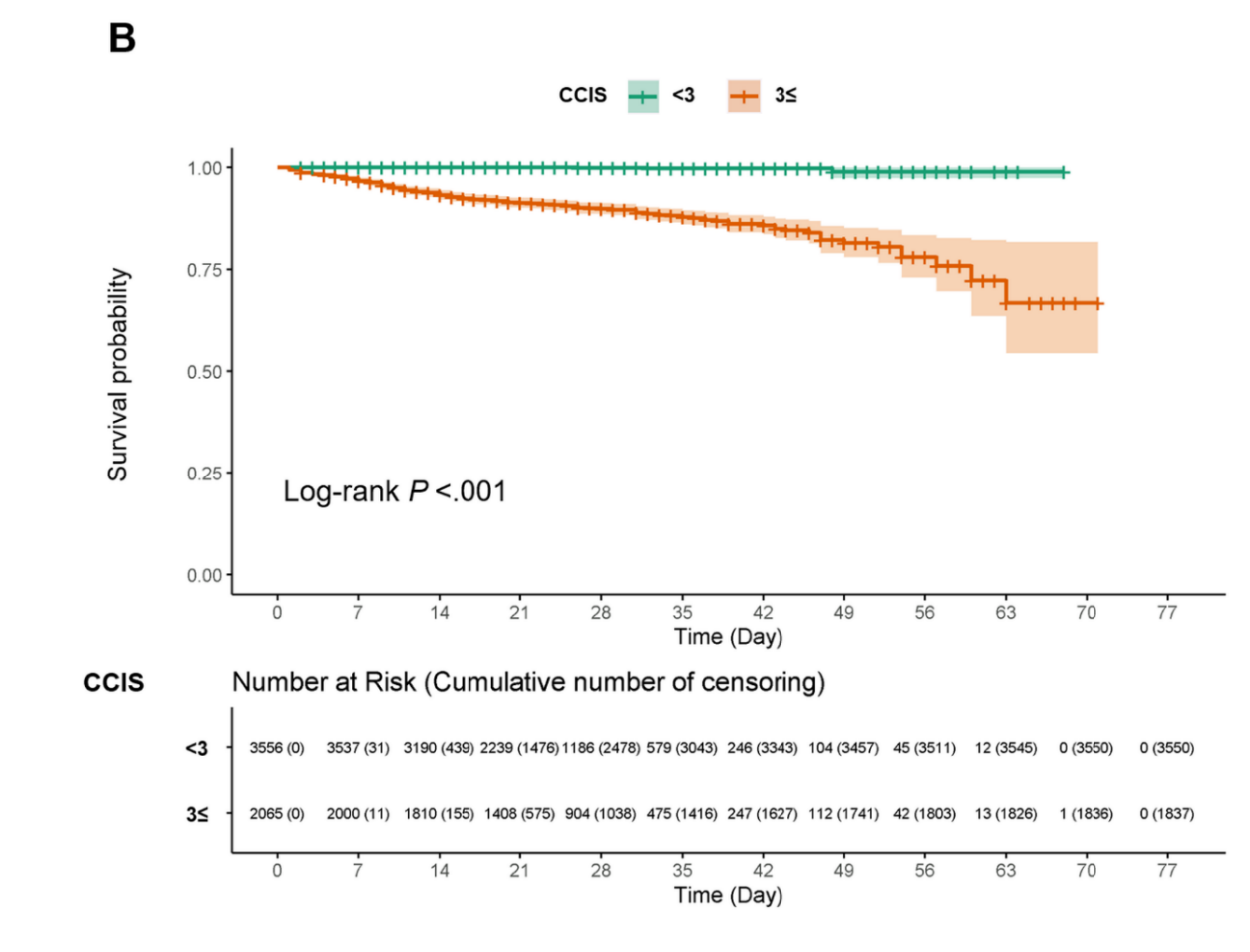

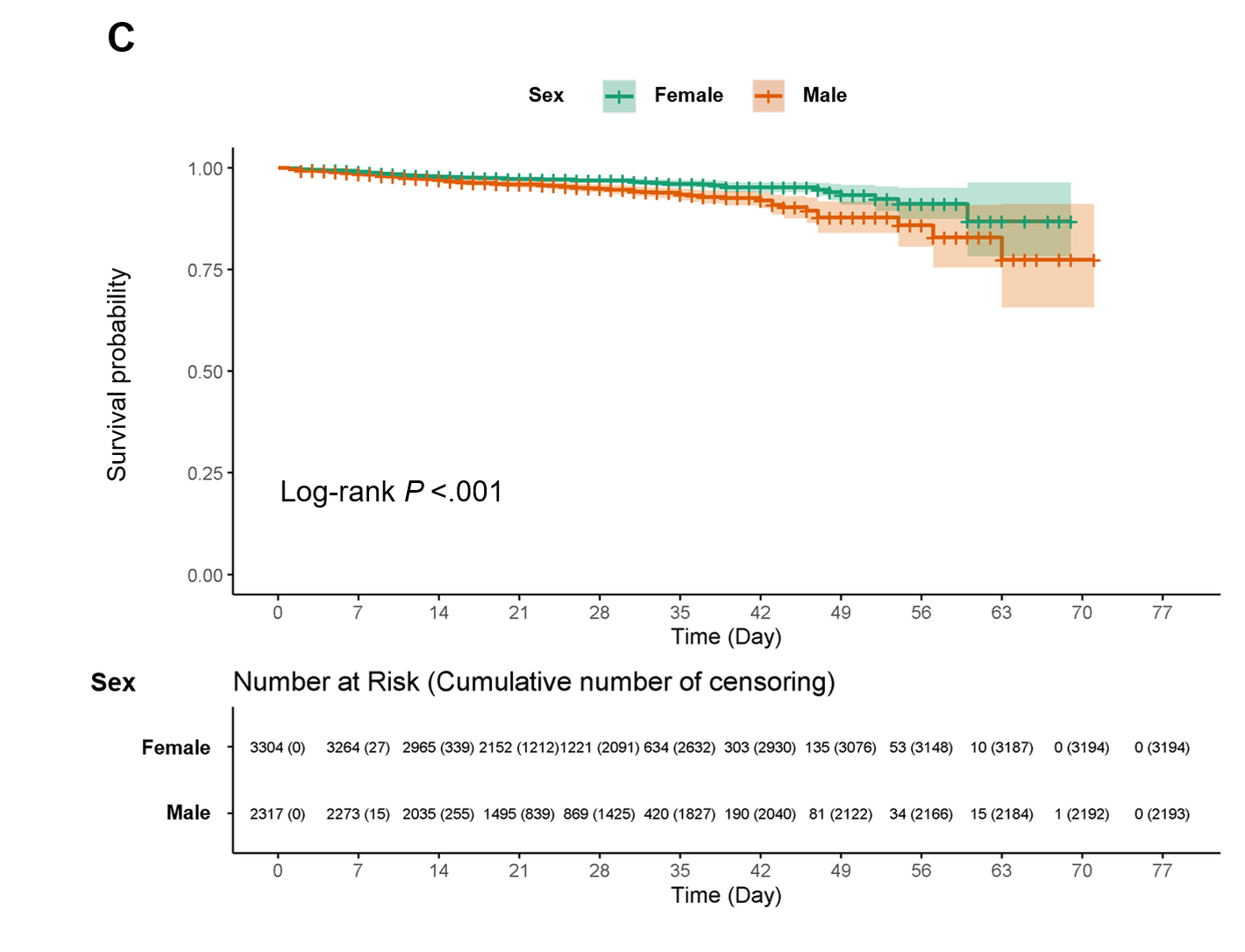

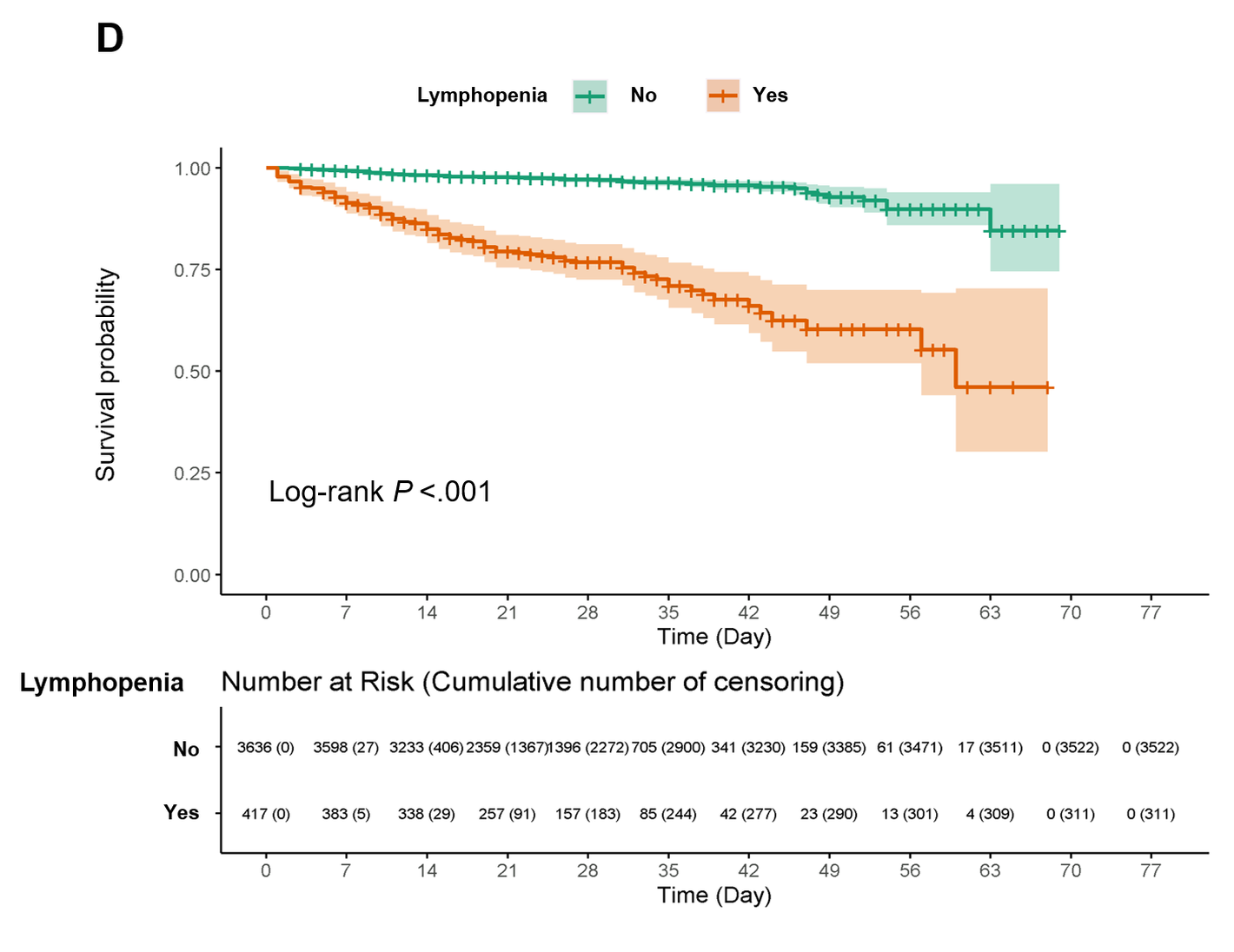

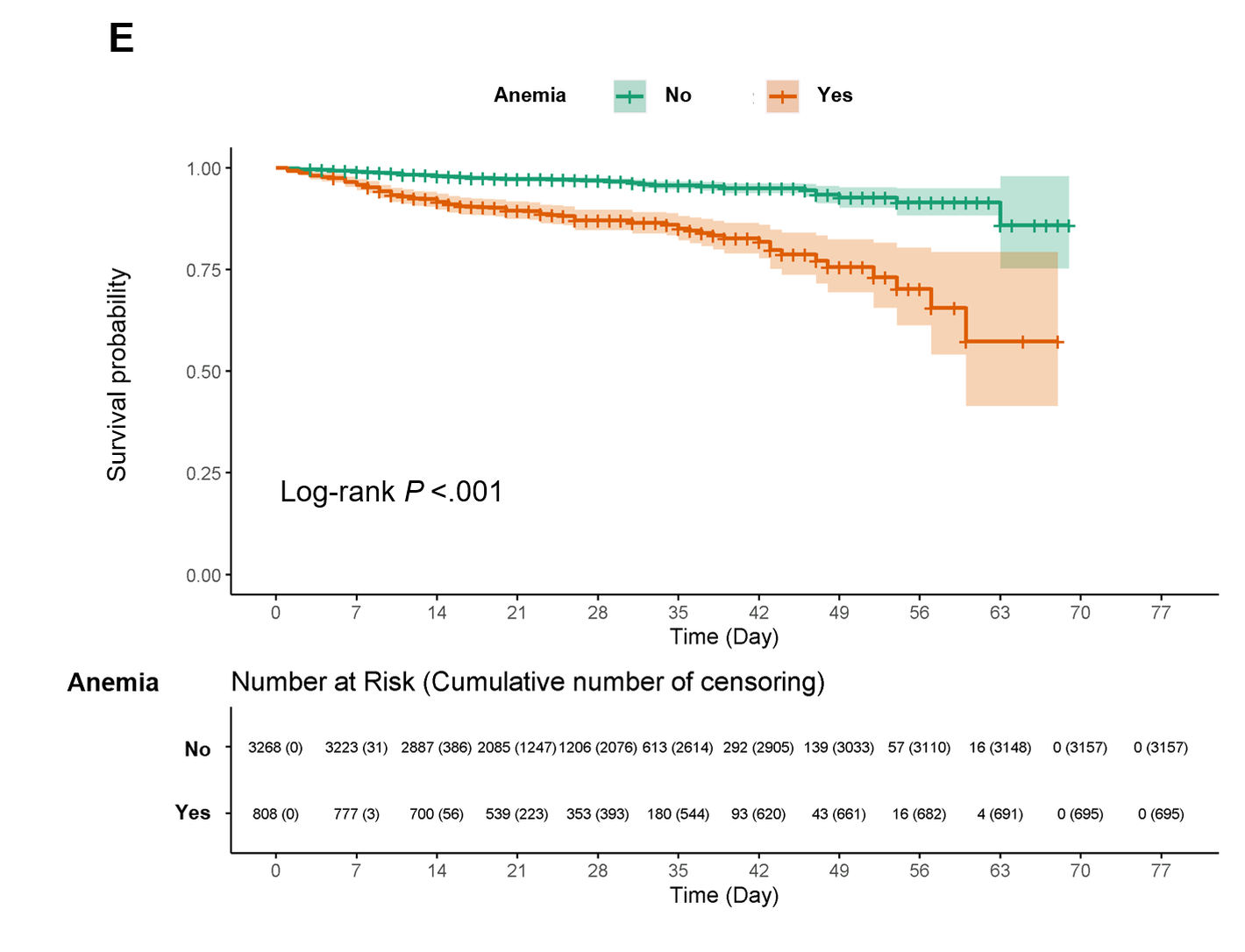

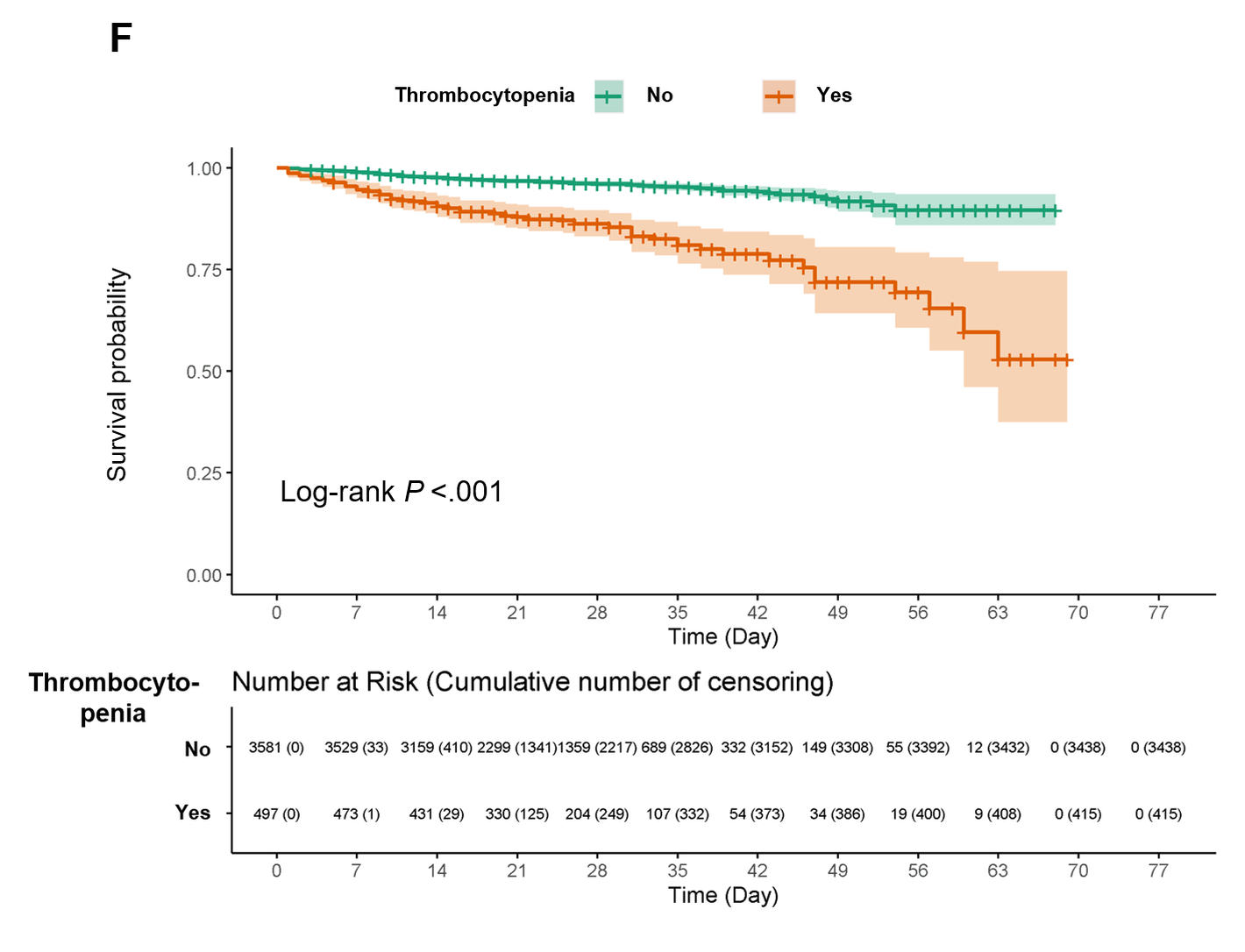

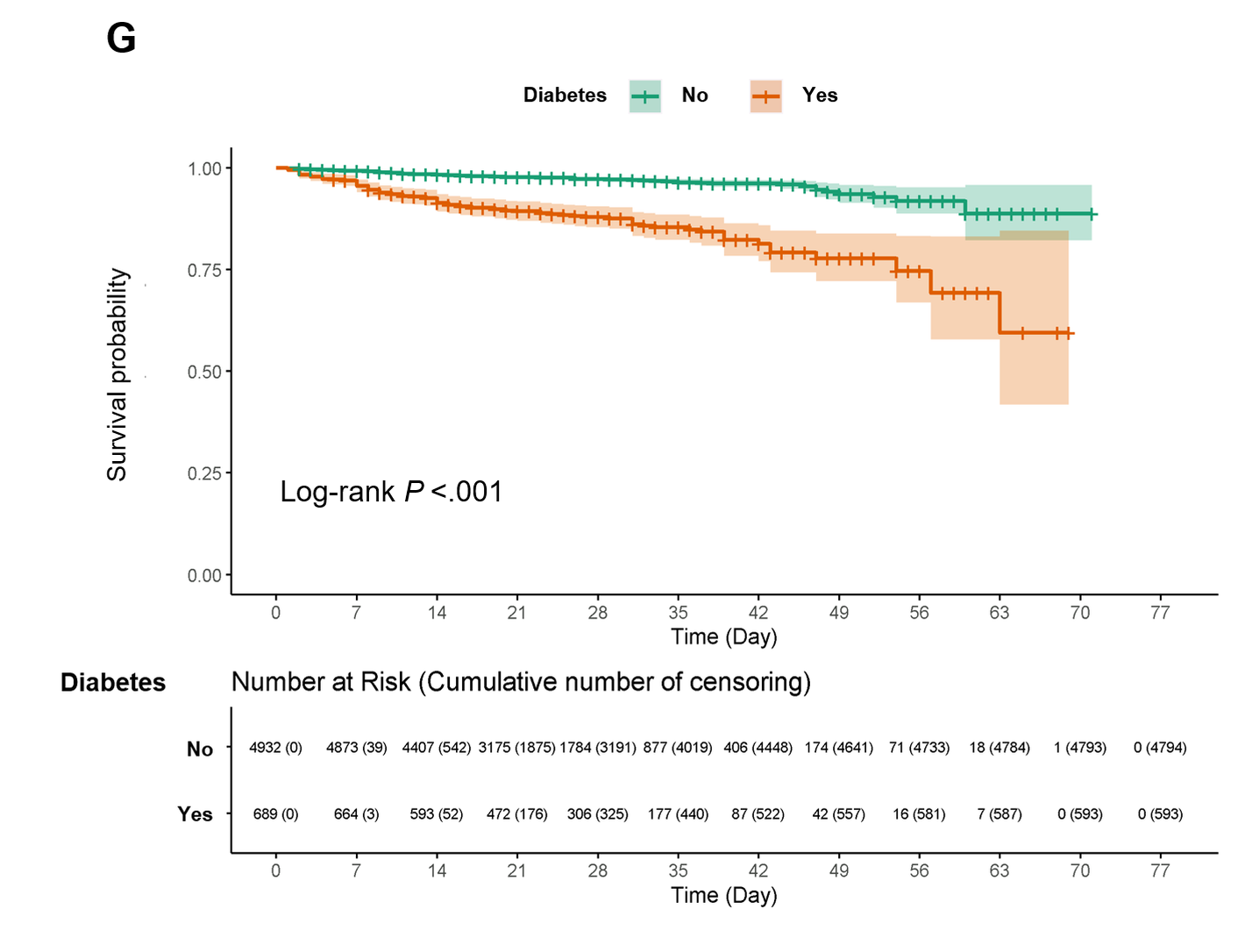

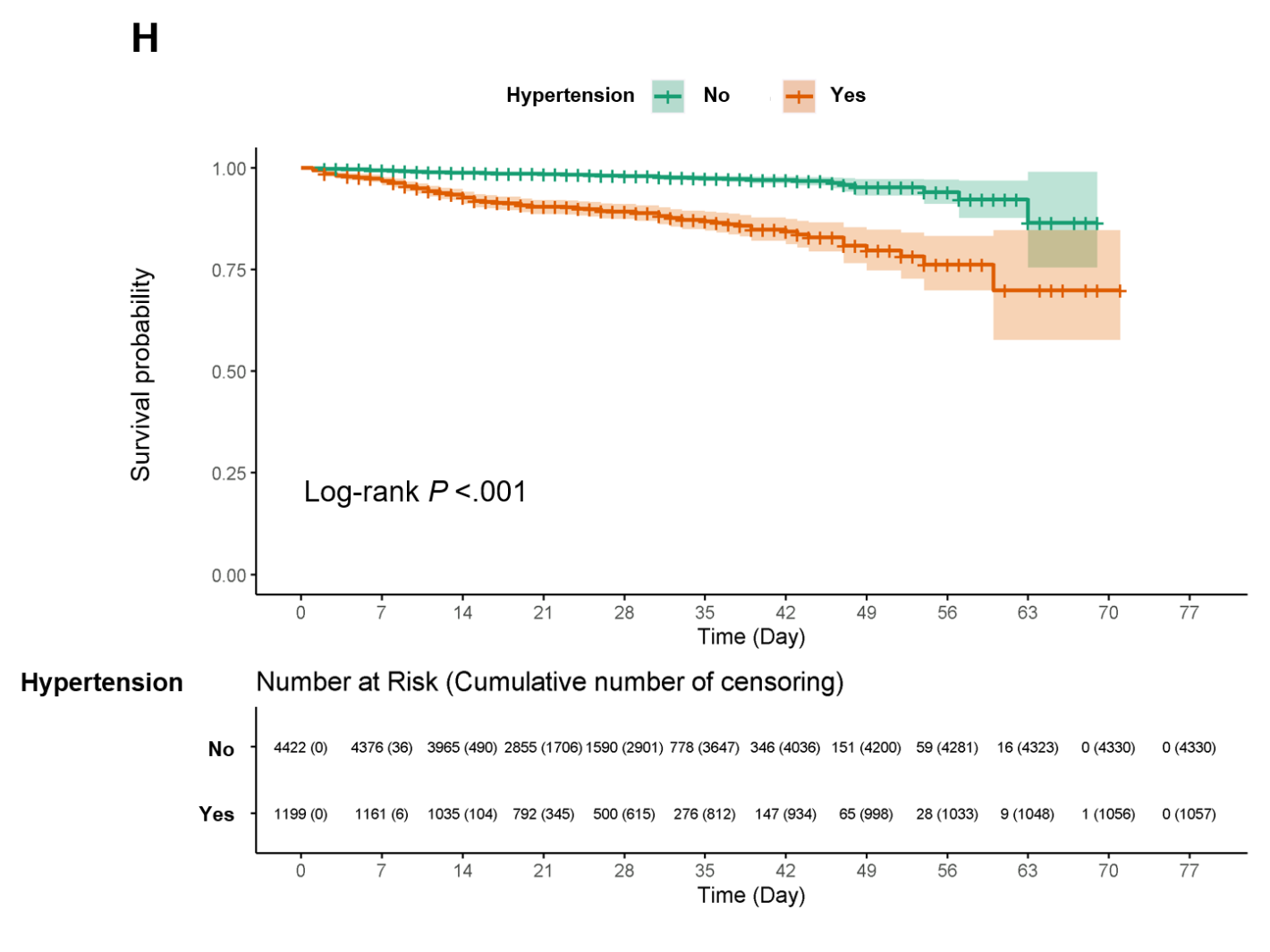

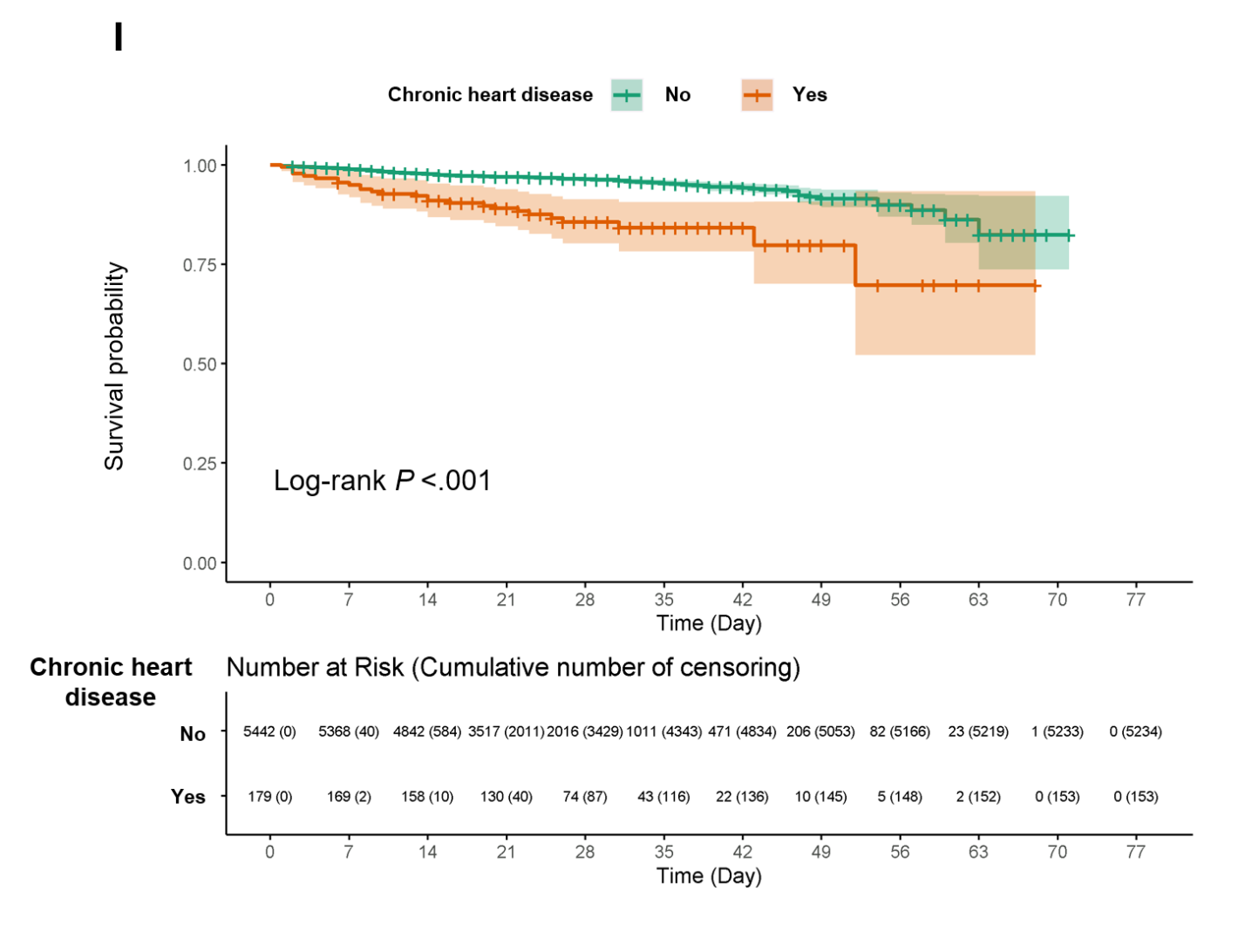

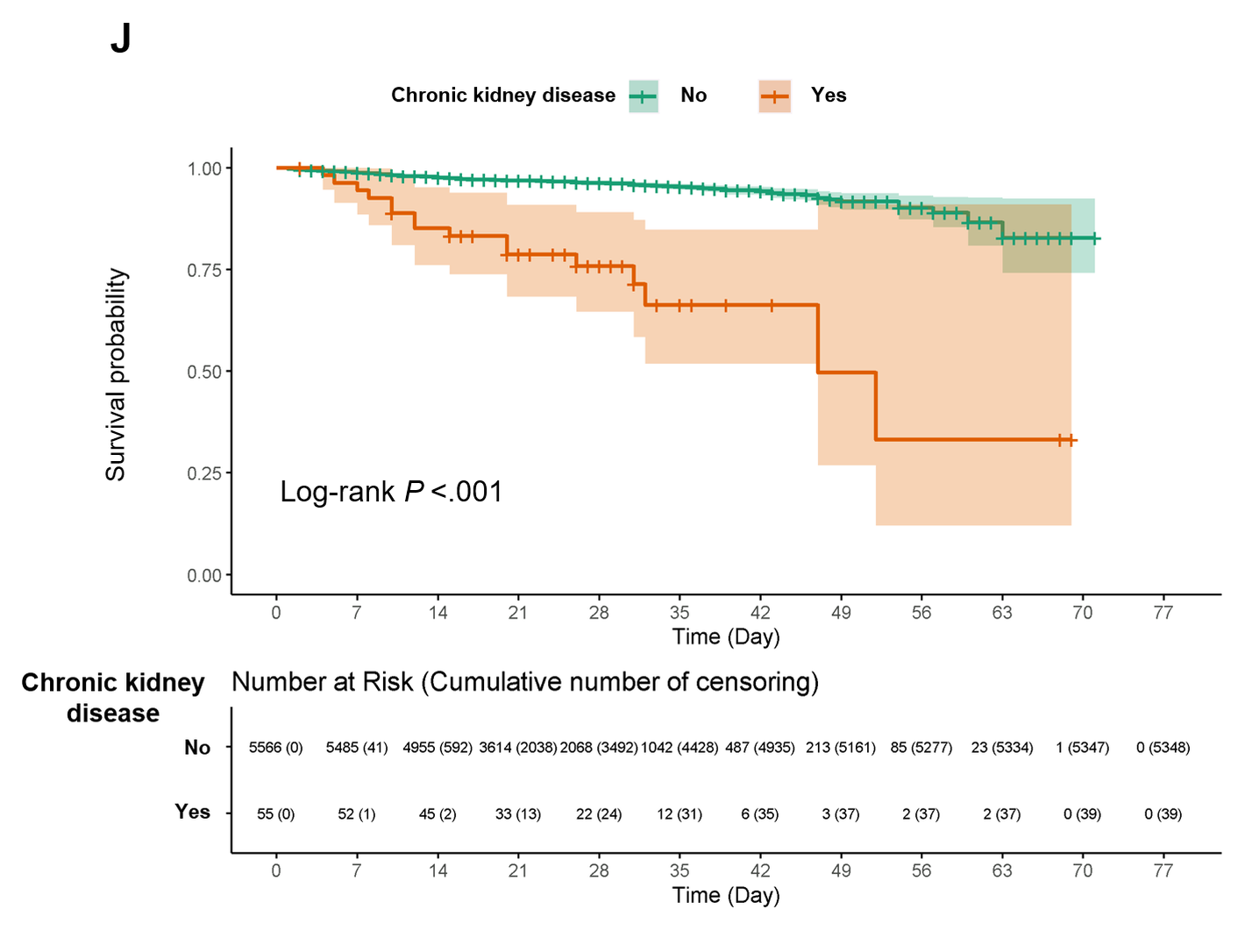

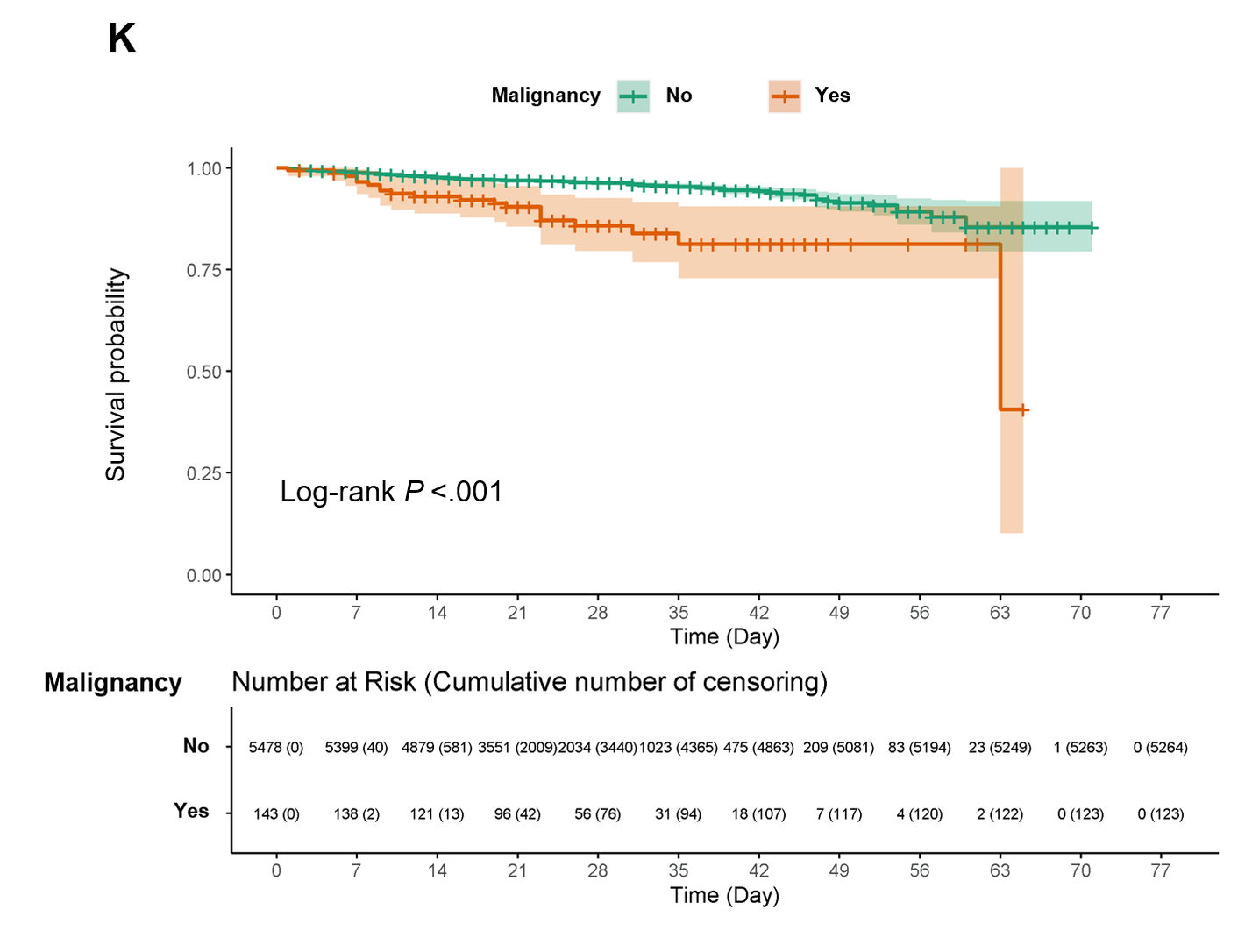

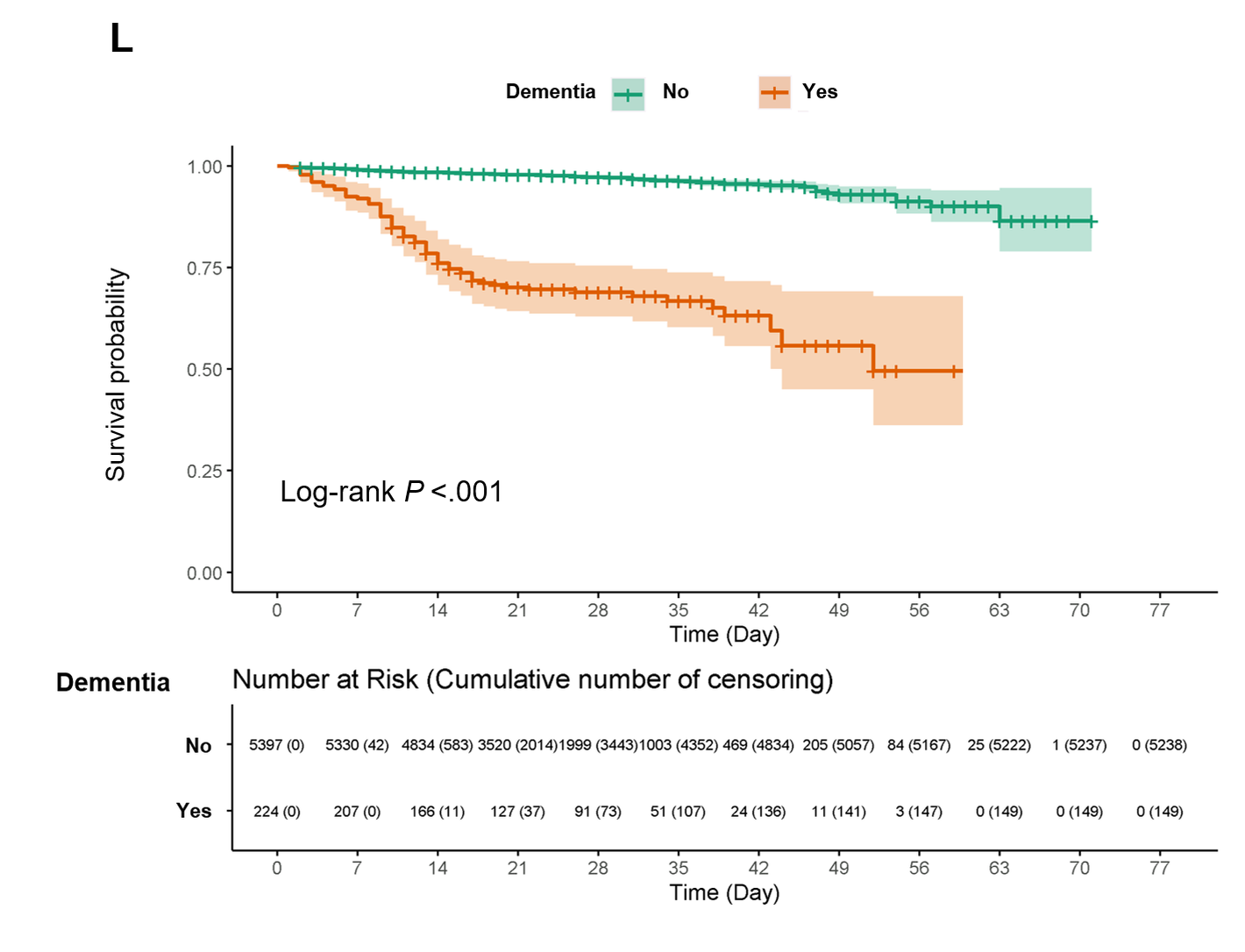

**Supplementary Figure S2. Calibration curves of the nomogram predicting 14-day (A), and 28-day (B) overall survival in patients with COVID-19.** We underwent bootstrap resampling 1,000 times to validate the nomogram. The C-index value for prediction of overall survival was 0.933, and R^2^ value was 0.99 in 14-day and 0.99 in 28-day prediction model. CCIS, age-adjusted Charlson comorbidity index score; COVID-19, coronavirus disease 2019.

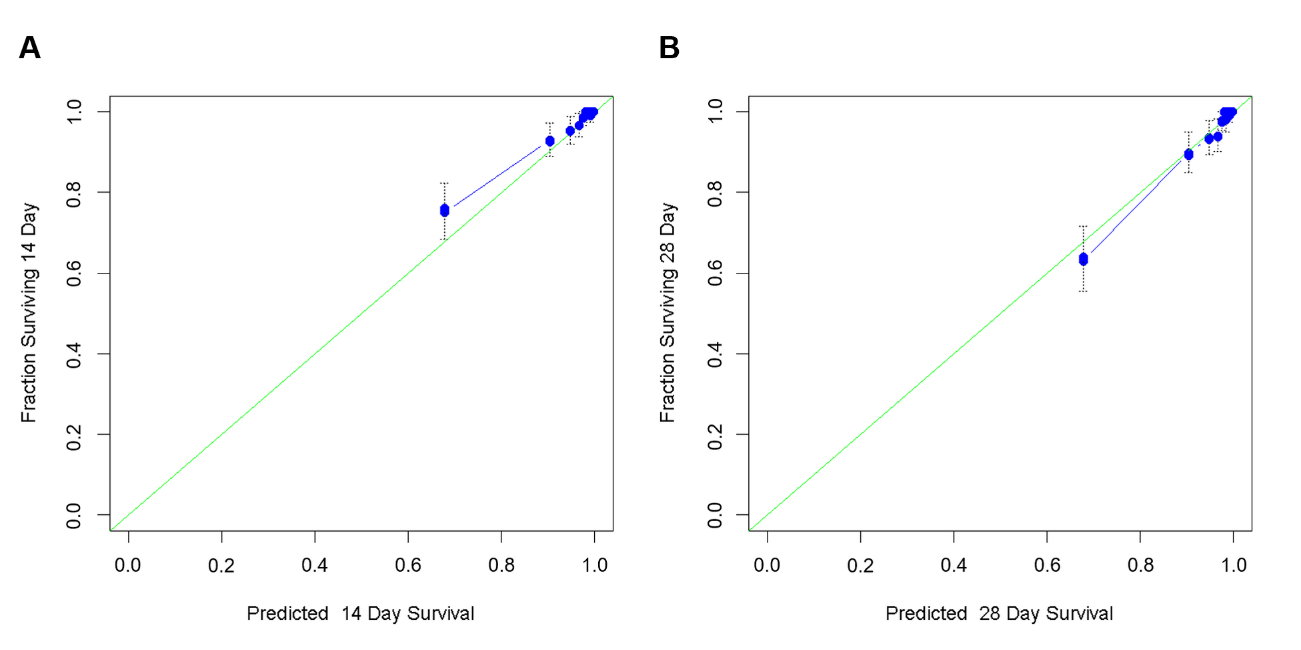
